# Clinical characteristics, outcomes and predictors of patients with leptospirosis admitted to medical intensive care unit: A retrospective review

**DOI:** 10.1101/2020.05.18.20105601

**Authors:** Atta Ajjimarungsi, Rungsun Bhurayanontachai, Sarunyou Chusri

## Abstract

**Background:** This study aimed to report the characteristics and outcomes of severe leptospirosis patients admitted to the ICU and to find the predictors of ICU admission.

**Methods:** This is a retrospective study of patients diagnosed as leptospirosis either from a serology diagnosis or Thai Lepto score (TLS) > 4. Patients were divided into ICU and general ward cases. All data were compared.

**Results:** Sixty-eight patients with severe leptospirosis were included. Thirty-two patients were ICU admission. ICU patients had higher SOFA score (9.8 ± 2.7 vs 4.6 ± 2.7; *p* = 0.001), higher TLS (7.6 ± 2.7 vs 5.7 ± 2.4; *p* < 0.001), lower hematocrit (32.7 ± 7.5% vs 36.6 ± 7.9%; *p* = 0.04), lower platelet count (88.7 ± 92.2 vs 173.3 ± 84.6 × 1000 cell/mm^3^; *p* = 0.008), higher total bilirubin (6.2 ± 6.3 vs 3.0 ± 3.6 mg/dL; *p* = 0.01), lower serum albumin (3.0 ± 0.5 vs 3.4 ± 0.7 g/dL; *p* = 0.03), higher creatinine (4.2 ± 2.4 vs 2.8 ± 2.1 mg/dL; *p* = 0.02), higher requirements for vasopressor or inotrope (90.6% vs 11.1%; *p* < 0.001), more mechanical ventilator support (84.4% vs 5.5%; *p* < 0.001), more renal replacement therapy (37.5% vs 0%; *p* < 0.001), higher hospital mortality (12.5% vs 0%; *p* = 0.03), and greater length of hospital stay (10.2 ± 5.1 vs 6.5 ± 5.1 days; *p* = 0.004). The TLS, mechanical ventilator support, and inotrope or vasopressor support were among the independent factors for ICU admission. A TLS > 6 indicated ICU admission.

**Conclusion:** The TLS, inotrope or vasopressor requirement, and mechanical ventilator support were the independent factors for ICU admission. TLS > 6 indicated ICU admission.

## 1. Introduction

Leptospirosis is a zoonotic disease of global importance[1, 2]. It has a worldwide distribution but is more common in the tropics where conditions for its transmission are particularly favorable[2]. In addition to the endemic burden, epidemics as well as massive outbreaks have been increasingly reported over the past decades[3-7]. Humans are usually infected through contact with water or soil contaminated with the urine of carrier animals[8]. Symptoms are usually flu-like including fever, headache, and myalgia and can resolve spontaneously. However, the disease can degenerate into severe forms evolving into multiple organ failure with hemorrhages and hepatic, renal, and pulmonary damage related to septic shock. Case fatality rates were reported to range from 5 to 70% depending on the clinical complications[9].

In this retrospective study, we aimed to evaluate the clinical characteristics and outcomes of hospitalized patients with a severe form of leptospirosis between the medical general ward and the medical intensive care unit in Songklanagarind Hospital during a period of 10 years and identify the factors influencing medical intensive care unit admission.

## 2. Methods

### 2.1 Patients

The electronic medical records of adult patients admitted with the final diagnosis of leptospirosis (ICD10-CM: A 27.0) between January 2006 and December 2016 at Songklanagarind Hospital were collected and reviewed. The study protocol was approved by the ethics committee at the Faculty of Medicine, Prince of Songkla University (EC number: 61-085-14-4). A waiver of consent was approved and the investigators agreed to confidentiality.

### 2.2 Data collection

All demographic data including gender, age, clinical presentations, duration of illness, history of water immersion, and season were recorded. Ward of admission was recorded as either general medical ward or medical intensive care unit (MICU).

The laboratory variables collected within the first 24 hours of hospital admission were complete blood count, international normalized ratio, blood urea nitrogen, creatinine, sodium, potassium, calcium, phosphate, creatine kinase, and liver function tests. Chest X-ray findings were also reviewed and classified into normal, focal infiltration, diffuse infiltration, and pleural effusion by the researcher (R.B). The Sequential Organ Failure Assessment score (SOFA) was then calculated from the biochemistry results.

The clinical parameters during hospitalization were also recorded including the results of the indirect immunofluorescence assay (IFA) for leptospirosis, Thai Lepto Score (TLS), arterial blood gas, type of antibiotics, time to antibiotics, acute lung injury index (PaO_2_/FiO_2_ ratio), and accumulative fluid balance. Artificial organ support was noted if the patients required mechanical ventilator support, renal replacement therapy, and used vasopressor or inotropic support. The treatment outcomes, including hospital length of stay, and hospital mortality were also recorded.

### 2.3 Definitions

After reviewing the selected medical records, the diagnoses of leptospirosis were classified into 2 types: definite cases and possible cases. The definite cases were identified if the patients were diagnosed as leptospirosis and the IFA titer for leptospirosis was greater than 1:400. The possible cases were patients diagnosed as leptospirosis with a negative IFA titer or no serological test for leptospirosis but had a TLS > 4.

The TLS is a tool for early diagnosis of leptospirosis which was recently developed and has been validated with good discrimination to diagnose clinically suspicious patients.[10] This score can be applied at the first visit while waiting for the serology results. Seven clinical and laboratory parameters (Table1) were integrated into the TLS. A composite score of 4 gave the AUROC value of 0.78 (95% confidence interval [CI] 0.68-0.89), sensitivity of 73.5, specificity of 73.7, positive predictive value of 87.8, and negative predictive value of 58.3.

**Table 1.** Thai Lepto score (TLS)

| **Parameters** | **Simplified Score** |
| --- | --- |
| **Hypotension (mean arterial blood pressure < 70 mmHg)** | 3 |
| **Jaundice (yellowish skin, sclera ad mucous membrane)** | 2 |
| **Muscle pain (non-traumatic muscle sore at calf and lower back)** | 2 |
| **Acute kidney injury according to ADIKO criteria** | 1.5 |
| **Hemoglobin < 12 gm/dL** | 3 |
| **Serum potassium < 3.5 mEq/L combined with serum sodium < 135 mEq/L** | 3 |
| **White blood cell count < 10,000/μL combined with PMN ≥ 80%** | 1 |

### 2.4 Statistical analysis

The continuous variables were expressed as mean and standard deviation (SD). The categorical variables were shown as number and percentage. To compare the clinical characteristics of the patients between general ward admission and MICU admission, the independent *t*-test or chi-square test was applied that depended on the type of variable. To identify the factors that influence MICU admission, the selected variables in univariable analysis with *p* < 0.2 were then introduced into a stepwise binary logistic regression model. All interested variables were tested for multicollinearity and were excluded from the model if a correlation was detected. The adjusted odds ratio (OR) and 95% CI were obtained and a *p*-value < 0.05 was statistically significant.

A receiver operating characteristic (ROC) curve and a calculated corresponding area under the ROC curve (AUROC) of TLS in the definite cases were subsequently constructed. The Youden index was introduced to select the best cut off value of TLS with the best sensitivity, specificity, positive likelihood ratio (LR+) and negative likelihood ratio (LR-) to predict the MICU admission. All statistical analyses used the MedCalc Statistical Software version 19.0.5 (MedCalc Software bvba, Ostend, Belgium; https://www.medcalc.org; 2019).

## 3. Results

### 3.1 Patient characteristics

Eighty-nine medical records with the final diagnosis of leptospirosis according to ICD-10 code were initially reviewed. Sixty-eight patients were included for the analysis. Forty-six (67.6%) patients were definite cases. Almost 80% presented during the wet season with the mean onset of symptoms of 5.3 ± 3.8 days. The five most common presentations in our cohort were fever (100%), myalgia (79.4%), jaundice (38.2%), chills (38.2%), and vomiting (36.8%). According to the disease severity and organ failure scores, patients had a mean SOFA score of 7 ± 4.1, and a mean TLS of 6.6 ± 2.7. From the biochemistry tests, we found that all cases had renal dysfunction with a mean creatinine of 3.5 ± 2.6 mg/dL and liver injury with a mean total bilirubin of 4.6 ± 5.3 mg/dL. Approximately 50% of cases had normal chest X-ray findings and 44.1% had diffuse lung infiltration. The most common antibiotics administered were ceftriaxone (75%) and doxycycline (50%). Almost half of the patients required inotropes or vasopressors and mechanical ventilator support. A minority of patients (17.6%) required renal replacement therapy. The overall survival rate was 94.1%. The demographic data, laboratory results, and treatment outcomes are presented in Table 2.

**Table 2.**
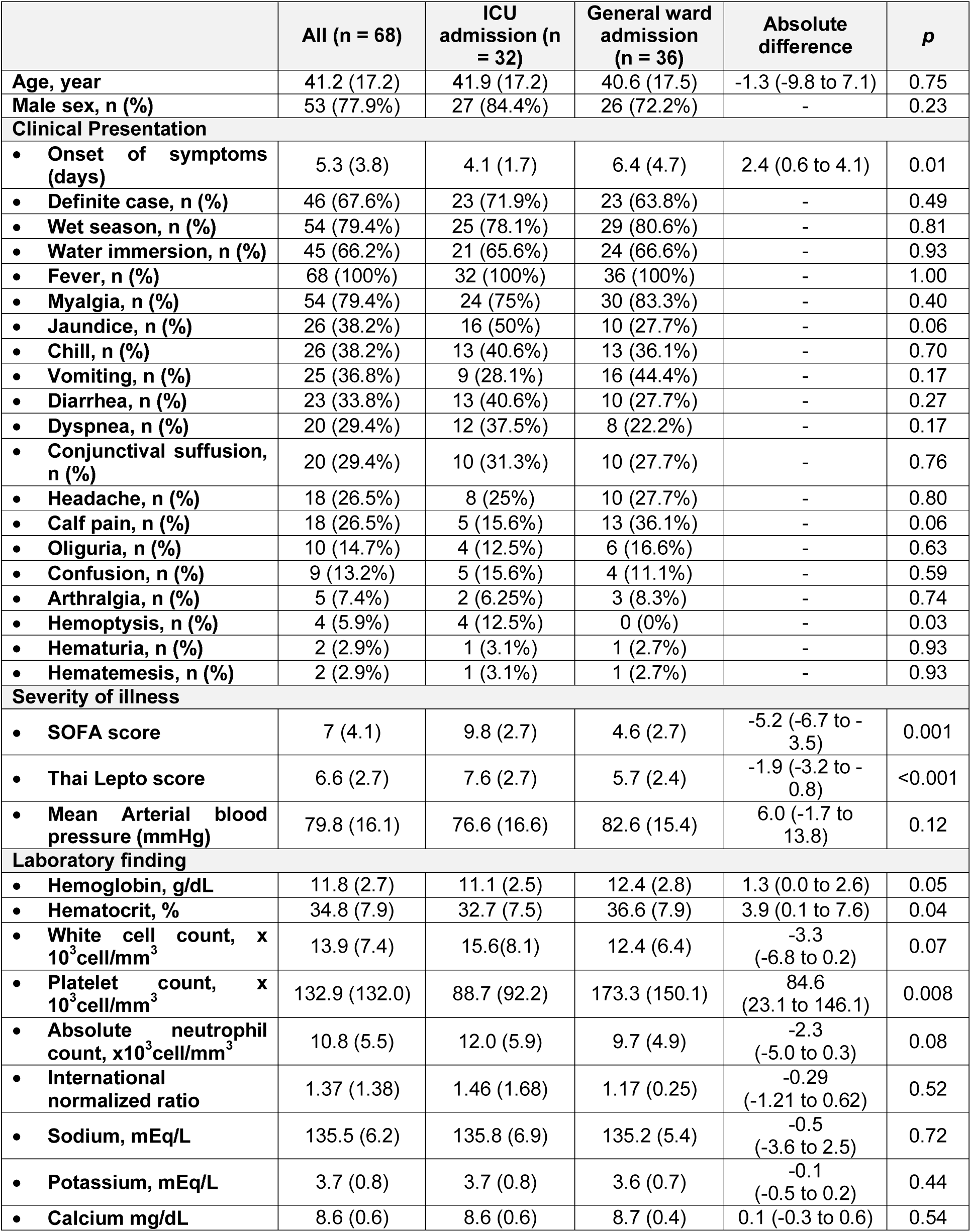
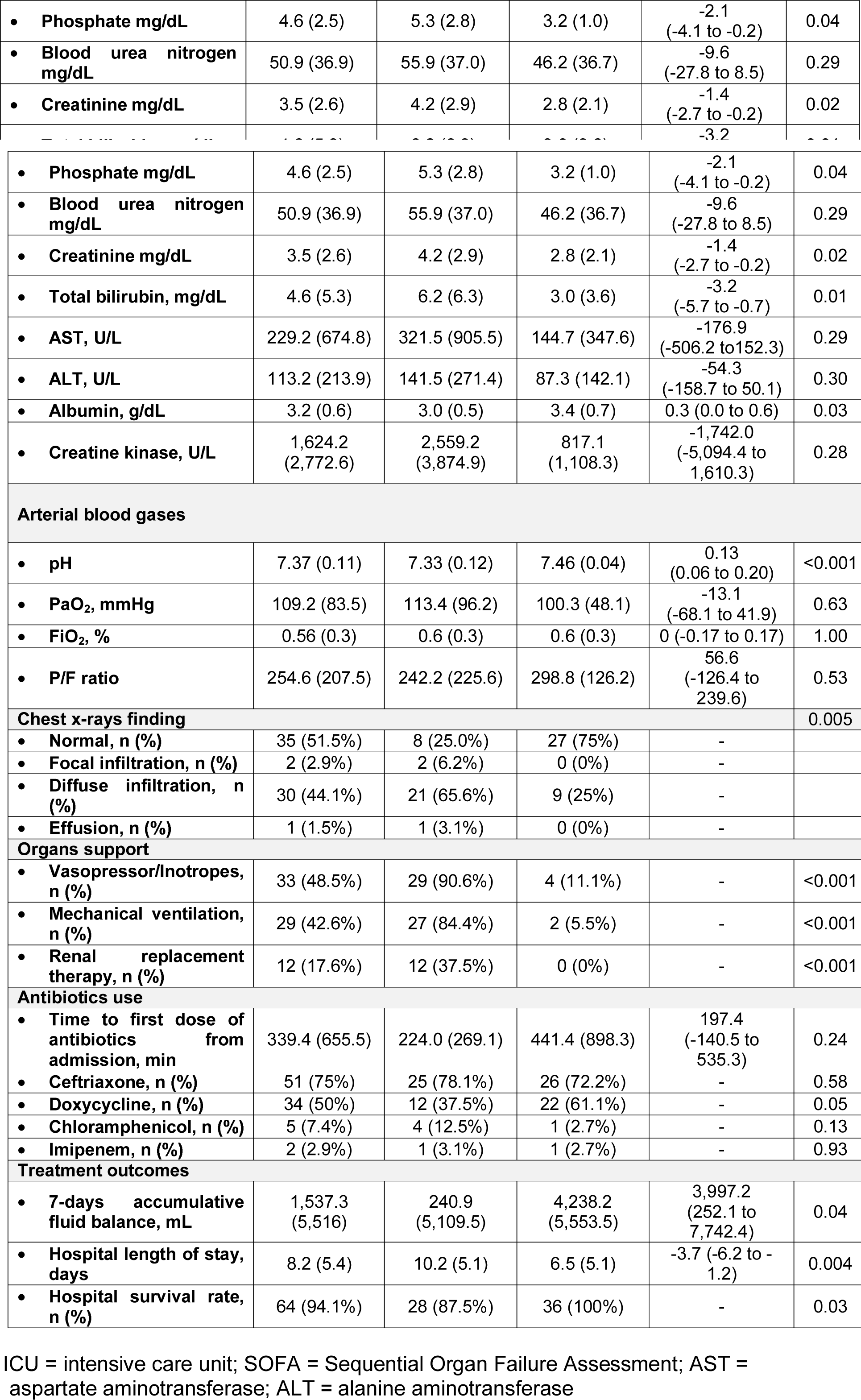
Clinical characteristics of severe leptospirosis patients between ICU and general ward admission.

|  | All (n = 68) | ICU admission (n = 32) | General ward admission (n = 36) | Absolute difference | p |
| --- | --- | --- | --- | --- | --- |
| Age, year | 41.2 (17.2) | 41.9 (17.2) | 40.6 (17.5) | -1.3 (-9.8 to 7.1) | 0.75 |
| Male sex, n (%) | 53 (77.9%) | 27 (84.4%) | 26 (72.2%) | - | 0.23 |
| **Clinical Presentation** |  |  |  |  |  |
| • Onset of symptoms (days) | 5.3 (3.8) | 4.1 (1.7) | 6.4 (4.7) | 2.4 (0.6 to 4.1) | 0.01 |
| • Definite case, n (%) | 46 (67.6%) | 23 (71.9%) | 23 (63.8%) | - | 0.49 |
| • Wet season, n (%) | 54 (79.4%) | 25 (78.1%) | 29 (80.6%) | - | 0.81 |
| • Water immersion, n (%) | 45 (66.2%) | 21 (65.6%) | 24 (66.6%) | - | 0.93 |
| • Fever, n (%) | 68 (100%) | 32 (100%) | 36 (100%) | - | 1.00 |
| • Myalgia, n (%) | 54 (79.4%) | 24 (75%) | 30 (83.3%) | - | 0.40 |
| • Jaundice, n (%) | 26 (38.2%) | 16 (50%) | 10 (27.7%) | - | 0.06 |
| • Chill, n (%) | 26 (38.2%) | 13 (40.6%) | 13 (36.1%) | - | 0.70 |
| • Vomiting, n (%) | 25 (36.8%) | 9 (28.1%) | 16 (44.4%) | - | 0.17 |
| • Diarrhea, n (%) | 23 (33.8%) | 13 (40.6%) | 10 (27.7%) | - | 0.27 |
| • Dyspnea, n (%) | 20 (29.4%) | 12 (37.5%) | 8 (22.2%) | - | 0.17 |
| • Conjunctival suffusion, n (%) | 20 (29.4%) | 10 (31.3%) | 10 (27.7%) | - | 0.76 |
| • Headache, n (%) | 18 (26.5%) | 8 (25%) | 10 (27.7%) | - | 0.80 |
| • Calf pain, n (%) | 18 (26.5%) | 5 (15.6%) | 13 (36.1%) | - | 0.06 |
| • Oliguria, n (%) | 10 (14.7%) | 4 (12.5%) | 6 (16.6%) | - | 0.63 |
| • Confusion, n (%) | 9 (13.2%) | 5 (15.6%) | 4 (11.1%) | - | 0.59 |
| • Arthralgia, n (%) | 5 (7.4%) | 2 (6.25%) | 3 (8.3%) | - | 0.74 |
| • Hemoptysis, n (%) | 4 (5.9%) | 4 (12.5%) | 0 (0%) | - | 0.03 |
| • Hematuria, n (%) | 2 (2.9%) | 1 (3.1%) | 1 (2.7%) | - | 0.93 |
| • Hematemesis, n (%) | 2 (2.9%) | 1 (3.1%) | 1 (2.7%) | - | 0.93 |
| **Severity of illness** |  |  |  |  |  |
| • SOFA score | 7 (4.1) | 9.8 (2.7) | 4.6 (2.7) | -5.2 (-6.7 to -3.5) | 0.001 |
| • Thai Lepto score | 6.6 (2.7) | 7.6 (2.7) | 5.7 (2.4) | -1.9 (-3.2 to -0.8) | <0.001 |
| • Mean Arterial blood pressure (mmHg) | 79.8 (16.1) | 76.6 (16.6) | 82.6 (15.4) | 6.0 (-1.7 to 13.8) | 0.12 |
| **Laboratory finding** |  |  |  |  |  |
| • Hemoglobin, g/dL | 11.8 (2.7) | 11.1 (2.5) | 12.4 (2.8) | 1.3 (0.0 to 2.6) | 0.05 |
| • Hematocrit, % | 34.8 (7.9) | 32.7 (7.5) | 36.6 (7.9) | 3.9 (0.1 to 7.6) | 0.04 |
| • White cell count, x 10<sup>3</sup>cell/mm<sup>3</sup> | 13.9 (7.4) | 15.6(8.1) | 12.4 (6.4) | -3.3 (-6.8 to 0.2) | 0.07 |
| • Platelet count, x 10<sup>3</sup>cell/mm<sup>3</sup> | 132.9 (132.0) | 88.7 (92.2) | 173.3 (150.1) | 84.6 (23.1 to 146.1) | 0.008 |
| • Absolute neutrophil count, x10<sup>3</sup>cell/mm<sup>3</sup> | 10.8 (5.5) | 12.0 (5.9) | 9.7 (4.9) | -2.3 (-5.0 to 0.3) | 0.08 |
| • International normalized ratio | 1.37 (1.38) | 1.46 (1.68) | 1.17 (0.25) | -0.29 (-1.21 to 0.62) | 0.52 |
| • Sodium, mEq/L | 135.5 (6.2) | 135.8 (6.9) | 135.2 (5.4) | -0.5 (-3.6 to 2.5) | 0.72 |
| • Potassium, mEq/L | 3.7 (0.8) | 3.7 (0.8) | 3.6 (0.7) | -0.1 (-0.5 to 0.2) | 0.44 |
| • Calcium mg/dL | 8.6 (0.6) | 8.6 (0.6) | 8.7 (0.4) | 0.1 (-0.3 to 0.6) | 0.54 |
| • Phosphate mg/dL | 4.6 (2.5) | 5.3 (2.8) | 3.2 (1.0) | -2.1<br>(-4.1 to -0.2) | 0.04 |
| • Blood urea nitrogen mg/dL | 50.9 (36.9) | 55.9 (37.0) | 46.2 (36.7) | -9.6<br>(-27.8 to 8.5) | 0.29 |
| • Creatinine mg/dL | 3.5 (2.6) | 4.2 (2.9) | 2.8 (2.1) | -1.4<br>(-2.7 to -0.2) | 0.02 |
| • Total bilirubin, mg/dL | 4.6 (5.3) | 6.2 (6.3) | 3.0 (3.6) | -3.2<br>(-5.7 to -0.7) | 0.01 |
| • AST, U/L | 229.2 (674.8) | 321.5 (905.5) | 144.7 (347.6) | -176.9<br>(-506.2 to 152.3) | 0.29 |
| • ALT, U/L | 113.2 (213.9) | 141.5 (271.4) | 87.3 (142.1) | -54.3<br>(-158.7 to 50.1) | 0.30 |
| • Albumin, g/dL | 3.2 (0.6) | 3.0 (0.5) | 3.4 (0.7) | 0.3 (0.0 to 0.6) | 0.03 |
| • Creatine kinase, U/L | 1,624.2<br>(2,772.6) | 2,559.2<br>(3,874.9) | 817.1<br>(1,108.3) | -1,742.0<br>(-5,094.4 to 1,610.3) | 0.28 |
| **Arterial blood gases** |  |  |  |  |  |
| • pH | 7.37 (0.11) | 7.33 (0.12) | 7.46 (0.04) | 0.13<br>(0.06 to 0.20) | <0.001 |
| • PaO<sub>2</sub>, mmHg | 109.2 (83.5) | 113.4 (96.2) | 100.3 (48.1) | -13.1<br>(-68.1 to 41.9) | 0.63 |
| • FiO<sub>2</sub>, % | 0.56 (0.3) | 0.6 (0.3) | 0.6 (0.3) | 0 (-0.17 to 0.17) | 1.00 |
| • P/F ratio | 254.6 (207.5) | 242.2 (225.6) | 298.8 (126.2) | 56.6<br>(-126.4 to 239.6) | 0.53 |
| **Chest x-rays finding** |  |  |  |  | 0.005 |
| • Normal, n (%) | 35 (51.5%) | 8 (25.0%) | 27 (75%) | - |  |
| • Focal infiltration, n (%) | 2 (2.9%) | 2 (6.2%) | 0 (0%) | - |  |
| • Diffuse infiltration, n (%) | 30 (44.1%) | 21 (65.6%) | 9 (25%) | - |  |
| • Effusion, n (%) | 1 (1.5%) | 1 (3.1%) | 0 (0%) | - |  |
| **Organs support** |  |  |  |  |  |
| • Vasopressor/Inotropes, n (%) | 33 (48.5%) | 29 (90.6%) | 4 (11.1%) | - | <0.001 |
| • Mechanical ventilation, n (%) | 29 (42.6%) | 27 (84.4%) | 2 (5.5%) | - | <0.001 |
| • Renal replacement therapy, n (%) | 12 (17.6%) | 12 (37.5%) | 0 (0%) | - | <0.001 |
| **Antibiotics use** |  |  |  |  |  |
| • Time to first dose of antibiotics from admission, min | 339.4 (655.5) | 224.0 (269.1) | 441.4 (898.3) | 197.4<br>(-140.5 to 535.3) | 0.24 |
| • Ceftriaxone, n (%) | 51 (75%) | 25 (78.1%) | 26 (72.2%) | - | 0.58 |
| • Doxycycline, n (%) | 34 (50%) | 12 (37.5%) | 22 (61.1%) | - | 0.05 |
| • Chloramphenicol, n (%) | 5 (7.4%) | 4 (12.5%) | 1 (2.7%) | - | 0.13 |
| • Imipenem, n (%) | 2 (2.9%) | 1 (3.1%) | 1 (2.7%) | - | 0.93 |
| **Treatment outcomes** |  |  |  |  |  |
| • 7-days accumulative fluid balance, mL | 1,537.3<br>(5,516) | 240.9<br>(5,109.5) | 4,238.2<br>(5,553.5) | 3,997.2<br>(252.1 to 7,742.4) | 0.04 |
| • Hospital length of stay, days | 8.2 (5.4) | 10.2 (5.1) | 6.5 (5.1) | -3.7 (-6.2 to -1.2) | 0.004 |
| • Hospital survival rate, n (%) | 64 (94.1%) | 28 (87.5%) | 36 (100%) | - | 0.03 |

ICU = intensive care unit; SOFA = Sequential Organ Failure Assessment; AST = aspartate aminotransferase; ALT = alanine aminotransferase

#### 3.2 Clinical characters between MICU admission and general medical ward admission with severe leptospirosis

Of 68 cases, 32 cases (47.06%) were admitted to the MICU and 20% of those had sepsis and septic shock. The MICU patients had a significantly shorter onset of symptoms (4.1 ± 1.7 vs 6.4 ± 4.7 days; *p* = 0.01), higher development of hemoptysis (12.5% vs 0%; *p* = 0.03), higher SOFA score (9.8 ± 2.7 vs 4.6 ± 2.7; *p* = 0.001), and higher TLS (7.6 ± 2.7 vs 5.7 ± 2.4; *p* < 0.001). From the blood results, the MICU patients had significantly lower hematocrit (32.7 ± 7.5% vs 36.6 ± 7.9%; *p* = 0.04), lower platelet count (88.7 ± 92.2 vs 173.3 ± 84.6 x 1000 cell/mm^3^; *p* = 0.008), higher serum creatinine level (4.2 ± 2.4 vs 2.8 ± 2.1 mg/dL; *p* = 0.02), higher phosphate level (5.3 ± 2.8 mg/dL vs 3.2 ± 1.0 mg/dL; *p* = 0.04), and higher total bilirubin level (6.2 ± 6.3 vs 3.0 ± 3.6 mg/dL; *p* = 0.01). On the other hand, the serum albumin was significantly lower in the ICU admission group (3.0 ± 0.5 vs 3.4 ± 0.7 g/dL; *p* = 0.03) and the chest X-ray results showed significantly greater infiltration in the MICU cases (66.6% vs 25%; *p* = 0.005).

Regarding organ support measurements and treatment outcomes, the MICU patients required more inotropes and vasopressors (90.6% vs 11.1%; *p* < 0.001), more mechanical ventilator support (84.4% vs 5.5%; *p* < 0.001), and more renal replacement therapy (37.5% vs 0%; *p* < 0.001). In addition, the MICU patients had a significantly longer length of hospital stay (10.2 ± 5.1 vs 6.5 ± 5.1 days; *p* = 0.004) and lower hospital survival (87.5% vs 100%; *p* = 0.03) compared to the general ward admission cases. The clinical characteristics and outcomes between MICU admission and general ward admission are shown in Table 2.

### 3.3 Factors related to MICU admission

Several interested parameters were excluded from the binary logistic regression analysis due to multicollinearity. We also found that the SOFA score was significantly correlated to the TLS with a correlation coefficient (r) of 0.6, *p* < 0.001. Therefore, the TLS was eventually selected for the calculated model. Finally, factors influencing MICU admission were TLS (OR 1.3; 95% CI 1.1-1.7, *p* < 0.001), requirement for mechanical ventilation (OR 63.2; 95% CI 5.8-691.4, *p* < 0.001), and use of inotropes and vasopressors (OR 53.5; 95% CI 5.5-524.5, *p* = 0.006).

After the selection of the definite cases of leptospirosis, a TLS > 6 could predict MICU admission with a sensitivity of 78.26%, specificity of 60.87%, positive likelihood ratio of 2.0, and negative likelihood ratio of 0.36 (AUROC 0.76, 95% CI 0.61-0.87), which was not inferior to SOFA score for predicting ICU admission (AUROC 0.88 vs 0.76; *p* = 0.05).

## 4. Discussion

Leptospirosis is a worldwide zoonotic infection commonly found in tropical areas around the world. The clinical presentation and clinical course have a wide range from mild symptoms to multiple organ dysfunction. Severe cases result in mortality and morbidity, which require MICU admission for artificial organ support. Our retrospective analysis reported the clinical characteristics of severe leptospirosis patients admitted into our hospital between 2006 and 2016. Our hospital is located in southern Thailand which is an endemic area for leptospirosis. Since our hospital is the biggest teaching and tertiary hospital in the area, the most severe patients are referred to us from the surrounding hospitals. From the results, we found that the patients with severe leptospirosis admitted to the MICU had higher disease severity and more patients required mechanical ventilator support, hemodynamic support, and renal replacement therapy. Almost 50% of the cases required MICU admission. Although the survival rate of the MICU cases was significantly lower than general ward admission, the overall survival rate of the severe form of leptospirosis in our cohort was quite high at 94.1%. We also found that either the TLS > 6 may indicate MICU admission.

In our study, we applied the TLS for the diagnosis and stratify the severity risk of leptospirosis. The TLS was established for the early diagnosis of leptospirosis while waiting for the serology results. This score consists of 7 parameters and a composite score greater than 4 gives a high sensitivity and specificity for the diagnosis of leptospirosis [[10]]. Although this score was initially intended for an early diagnosis, we also found a significant correlation with the SOFA score. Therefore, the TLS could possibly be used as a severity index to triage patients to the appropriate care area. We found that a TLS > 6 gives good discriminating power for ICU admission with an AUROC of 0.76 (95% CI: 0.61-0.87). Furthermore, this composite score is an independent factor for MICU admission with an OR of 1.3 (95% CI: 1.1-1.7, *p* < 0.001) as well as mechanical ventilator support and inotrope or vasopressor requirements.

A recent study from Brazil demonstrated that tachypnea, hypotension, and acute kidney injury were among the independent factors for ICU admission[11] and the results were similar to our findings. That study also found that a significant number of elderly patients were admitted to the ICU. Also, a study from India indicated that aging was a risk factor for ICU admission[12]. Unfortunately, our study did not support those findings. The mean age of both MICU admission and general ward admission was lower than the previous studies and was not significantly different between the groups.

Although several studies indicated that pulmonary and kidney dysfunction in leptospirosis was significantly related to mortality and bad clinical outcomes[11- 13], our study reported a lower mortality rate of 5.9% compared to those previous studies. Our cohort demonstrated kidney and lung injury in both the MICU and general ward cases. In all patients (n = 68) the mean serum creatinine level was 3.5 ± 2.6 mg/dL and mild lung injury had a mean P/F ratio of 254.6 ± 207.5. The reason for the lower mortality rate of severe leptospirosis in our cohort could be from the younger age and most of the cases received antibiotics as either ceftriaxone or doxycycline. Several studies demonstrated that age is a risk factor in both ICU admission and mortality[11, 12, 14]. Furthermore, antibiotic treatment, including ceftriaxone, was a preventive factor for ICU admission and mortality in some studies[15, 16]. Although there was no consensus for antibiotics in leptospirosis, several experts have recommended starting antibiotics such as doxycycline or ceftriaxone in severe cases who present with multiple organ failure[17, 18].

Our study reported the most severe cases of leptospirosis that developed multiple organ involvement including liver injury, kidney injury, lung injury, and hemodynamic instability. Almost half of the patients required organ support including inotropes or vasopressors, mechanical ventilation, and renal replacement therapy. In resource-limited settings without serological evidence or a lack of time to obtain a serological diagnosis of leptospirosis, we suggest using the TLS not only for the early diagnosis but for severity assessment to allocate the patients to the most appropriated care area. A patient with severe leptospirosis and a TLS score greater than 6 should be admitted to the ICU for optimal management with organ support and antibiotics.

However, our study had several limitations. First, the study was retrospective in nature and some variables were missing or unrecorded which possibly led to study bias. In addition, the results of our study may not be applied for less severe leptospirosis cases or may not represent the general population infected with a mild self-limiting form of leptospirosis. Last, the definite cases of leptospirosis with positive serology test in our study was only 67.6%, the other possible cases were possibly infected by other organisms that responded to the optimal antibiotic management. However, we corrected that defect by applying the highly sensitive and specific composite score for the diagnosis of leptospirosis. In addition, the definite cases in both groups were not significantly different.

In conclusion, mechanical ventilator support and inotrope or vasopressor requirements were among the independent factors for ICU admission in the severe form of leptospirosis. The TLS or SOFA score could be applied at the time of admission to triage the patients to the ICU care. A TLS > 6 indicated ICU admission in severe leptospirosis.

## Data Availability

Not applicable

## Acknowledgements

The authors would like to gratefully thanks the native English speaker mentor from the International Affair Unit, Faculty of Medicine, Prince of Songkla University for the language correction service.

## Funding support

This study is funded by the research fund, Faculty of Medicine, Prince of Songkla University, Hat Yai, Songkhla, Thailand.

## Financial Disclosure

None declared

## Conflict of interest statement

All authors declared no conflicts of interest.

## Listing of all authors

Atta Ajjimarungsi, Department of Medicine, Faculty of Medicine, Prince of Songkla University, Hat Yai, Songkhla, Thailand. Rungsun Bhurayanontachai, Division of Critical Care Medicine, Department of Medicine, Faculty of Medicine, Prince of Songkla University, Hat Yai, Songkhla, Thailand. Sarunyou Chusri, Division of Infectious disease, Department of Internal Medicine, Faculty of Medicine, Prince of Songkla University, Songkhla, Thailand.

## Authorship statement

Atta Ajjimarungsi, Rungsun Bhurayanontachai and Sarunyou Chusri equally contributed to the conception/design of the research. Atta Ajjimarungsi, Rungsun Bhurayanontachai contributed to the acquisition, analysis, and interpretation of the data, and drafted the manuscript; Rungsun Bhurayanontachai critically revised the manuscript; and all authors agree to be fully accountable for ensuring the integrity and accuracy of the work and read and approved the final manuscript

## Notes

### Competing Interest Statement

The authors have declared no competing interest.

## References

1. Vinetz JM: Leptospirosis. Current opinion in infectious diseases 2001, 14(5):527–538.

2. Bharti AR, Nally JE, Ricaldi JN, Matthias MA, Diaz MM, Lovett MA, Levett PN, Gilman RH, Willig MR, Gotuzzo E et al: Leptospirosis: a zoonotic disease of global importance. The Lancet Infectious Diseases 2003, 3(12):757–771.

3. Agampodi SB PS, Thevanesam V, Nugegoda DB, Smythe L, Thaipadungpanit J et al Leptospirosis outbreak in Sri Lanka in 2008: lessons for assessing the global burden of disease.. Am J Trop Med Hyg 85(3):471–478 2011.

4. Amilasan AS UM, Suzuki M, Salva E, Belo MCP, Koizumi N et al (2012).: Outbreak of leptospirosis after flood, the Philippines, 2009. Emerging infectious diseases 2012, 18(1):91–94.

5. Barcellos C SP: The place behind the case: leptospirosis risks and associated environmental conditions in a flood-relatedoutbreak in Rio de Janeiro. Cad Saude Publica 2001, 17(Suppl):59–67.

6. Cruz LS VR, Lopes AA (2009).: Leptospirosis: a worldwide resurgent zoonosis and important cause of acute renal failure and death in developing nations. Ethn Dis 2009, 19(1 Suppl 1):37–41.:37-41.

7. Goarant C L-BS, Perez J, Vernel-Pauillac F, Chanteau S, Guigon A Outbreak of leptospirosis in New Caledonia: diagnosis issues and burden of disease. Trop Med Int Health 2009, 14(8):926–929.

8. Hochedez P, Theodose R, Olive C, Bourhy P, Hurtrel G, Vignier N, Mehdaoui H, Valentino R, Martinez R, Delord JM et al: Factors Associated with Severe Leptospirosis, Martinique, 2010-2013. Emerg Infect Dis 2015, 21(12):2221- 2224.

9. Mikulski M, Boisier P, Lacassin F, Soupe-Gilbert ME, Mauron C, Bruyere-Ostells L, Bonte D, Barguil Y, Gourinat AC, Matsui M et al: Severity markers in severe leptospirosis: a cohort study. Eur J Clin Microbiol Infect Dis 2015, 34(4):687–695.

10. Sukmark T, Lumlertgul N, Peerapornratana S, Khositrangsikun K, Tungsanga K, Sitprija V, Srisawat N, Thai-Lepto AKIsg: Thai-Lepto-on-admission probability (THAI-LEPTO) score as an early tool for initial diagnosis of leptospirosis: Result from Thai-Lepto AKI study group. PLoS Negl Trop Dis 2018, 12(3):e0006319.

11. Daher Ede F, Soares DS, de Menezes Fernandes AT, Girao MM, Sidrim PR, Pereira ED, Rocha NA, da Silva GB, Jr.: Risk factors for intensive care unit admission in patients with severe leptospirosis: a comparative study according to patients’ severity. BMC Infect Dis 2016, 16: 40.

12. Goswami RP, Goswami RP, Basu A, Tripathi SK, Chakrabarti S, Chattopadhyay I: Predictors of mortality in leptospirosis: an observational study from two hospitals in Kolkata, eastern India. Trans R Soc Trop Med Hyg 2014, 108(12):791–796.

13. Cerqueira TB, Athanazio DA, Spichler AS, Seguro AC: Renal involvement in leptospirosis--new insights into pathophysiology and treatment. Braz J Infect Dis 2008, 12(3):248–252.

14. Panaphut T, Domrongkitchaiporn S, Thinkamrop B: Prognostic factors of death in leptospirosis: a prospective cohort study in Khon Kaen, Thailand. Int J Infect Dis 2002, 6(1):52–59.

15. Faucher JF, Chirouze C, Hoen B, Leroy J, Hustache-Mathieu L, Estavoyer JM: Short-course treatment with ceftriaxone for leptospirosis: a retrospective study in a single center in Eastern France. J Infect Chemother 2015, 21(3):227–228.

16. Panaphut T, Domrongkitchaiporn S, Vibhagool A, Thinkamrop B, Susaengrat W: Ceftriaxone compared with sodium penicillin g for treatment of severe leptospirosis. Clin Infect Dis 2003, 36(12):1507–1513.

17. Raptis L, Pappas G, Akritidis N: Use of ceftriaxone in patients with severe leptospirosis. Int J Antimicrob Agents 2006, 28(3):259–261.

18. Faucher JF, Hoen B, Estavoyer JM: The management of leptospirosis. Expert Opin Pharmacother 2004, 5(4):819–827.

